# Enhancing Palliative Care Registration Using an Automated Robotic Process with Clinician Validation: A PRADA Prospective Cohort Study

**DOI:** 10.1101/2025.08.27.25334555

**Authors:** Emily Heyting, Baldev Singh, Vijay Klaire, Nisha Kumari, Kam Ahmed, John Burrell, Elizabeth Guest, Abdool Koodaruth, Prasadika Labutale, Khalid Mahmood, Gillian Pickavance, Mona Sidhu, Gurjeet Malhi, Emma Parry

**Author notes:** **<u>Corresponding author</u>** PARRY Emma, NIHR Clinical Lecturer in Primary Care. **<u>Author contributions</u>** Principal authors EH, EP, BMS. Wolverhampton Digital Health Care Primary Care Research Network KA, JB, EG, AK, PL, GM, KM, GP, MS. Data systems VK, NK, JL. Authors with full access to data BMS VK EP. Statistical Analysis BMS. Manuscript development, all.

## Abstract

**Background:** Ineffective care planning at the end of life carries high rates of unscheduled hospitalisation and avoidable deaths in hospital. Palliative care registration (PCR) and crucially, care planning, helps improve adverse outcomes. However, identification for registration often occurs late or not at all.

**Aim:** To improve PCR accuracy and uptake by developing an automated digital tool, utilising palliative care process markers, combined with clinical judgement, to identify those with palliative care needs.

**Design and Setting:** A primary care based prospective cohort study in Wolverhampton.

**Method:** All adults (n=39,079) were included. A robotic process tool (BOT) identified those not on the PCR using any of five palliative care process markers. BOT positive patients were assessed by GPs for the PCR. Performance metrics for prediction of subsequent 1 year mortality were determined.

**Results:** 546 patients were BOT positive. GPs added 131 patients (24%) to the PCR. This subset had the highest mortality rate (48.1%) compared to; those already on the PCR (37.2%), BOT positive patient who were not added to the PCR (19.5%) and those who were non-registered and BOT negative (0.8%) (p<0.001). The new combined PCR captured 220 (35.5%) of deaths, compared to 25.4% in the original PCR. A ‘digital safety net’ group, comprising those initially on the PCR and all BOT positive patients, accounted for 48.6% of deaths.

**Conclusion:** We developed a robotic process technique combined with GP assessment that yielded higher PCR uptake and accuracy, identifying individuals at-risk for ongoing surveillance.

**How this fits in:** Palliative care registers support the delivery of good palliative care by ensuring those who may be nearing the end of life have an assessment of their needs and supported care planning. Previous tools to support identification for palliative care registration have relied on opportunistic identification. Many people who die expectedly are not included on registers. Our new method demonstrates an automated, electronic, data driven system incorporating clinician validation. This system improved the uptake and accuracy of palliative care registration.

## Introduction

Increasing clinical complexity and aging populations strain health services (1). Patients nearing the end-of-life experience high rates of inappropriate admissions and death in hospital (2). Amongst extensive guidance for better palliative care, earlier identification of those approaching the end-of-life for palliative care registration (PCR) is paramount (1,3,4). PCR was part of the UK NHS primary care Quality and Outcome Framework (QOF) until retired in 2025, although it remains as part of NHS business processes. Inclusion on a PCR promotes care coordination, increases advanced care planning and improves the likelihood of dying in a preferred place (5) but only 55% of English practices established and maintained a palliative care register in 2024-2025 (6). It is estimated that 75% of deaths are potentially predictable, however less than 30% of those who die are captured by PCR (7,8). General practitioners (GPs) find the workload of maintaining PCR valuable but time consuming (9). This calls for new ways of working.

For early identification, the Surprise Question for clinician defined mortality prognosis is of uncertain validity and does not consistently perform well at predicting death (10). Several tools aid identification of patients with palliative care needs, but these have been developed in small populations, in disease specific cohorts or are not automated for application at scale (11–14). Addressing the failure of identification for PCR requires a systematic approach with modern day digital heath techniques (15). Methodologies for advanced data engines are still emergent (16) and complexity in algorithmic approaches with “big data” and machine learning may not confer improved metrics in health prediction or outcomes (17).

An uncomplicated digital system that supports establishing, maintaining, and reviewing the palliative care register, and which provides standardised performance metrics and evidenced outcomes is urgently needed. We have integrated data from hospital, community and primary care sources and deployed this into direct care as a clinical governance, quality improvement and clinical decision support tool. This tool provides early identification for anticipatory care, capturing required processes for structured care and ascertaining prospective events to determine effectiveness. We have termed this ‘The Proactive Risk Based and Data Driven Assessment of Patients at the End of Life’ (PRADA). Our published evidence outlines its general methodology, utility and its potential for benefit (18–20).

We have now developed an automated robotic process tool to improve PCR accuracy and uptake, utilising end of life process markers to escalate patients to their GP who makes an assessment regarding inclusion on the PCR. We emphasise the distinct but arising opportunity for needs assessment and care planning during this process, however this was not the focus of this study. Specifically, we evidence the system’s ability to improve PCR accuracy and uptake, validating the assessment outcomes against 1 year mortality.

## Methods

### Study setting

Eight general practices in Wolverhampton, a UK multiethnic, high deprivation, industrial city.

### Study design

We selected the whole live adult population (age ≥ 18 years, n=39, 079) of included practices. No exclusion criteria were applied. Patients were characterised according to their PCR status. Five process markers were ascertained from our electronic, integrated hospital, community, and primary care records: the Surprise Question outcome “No”, Electronic Palliative Care Coordinating System documentation, Recommended Summary Plan for Emergency Treatment and Care documentation, any advanced care plans and any coded contact with the hospital Specialist Palliative Care team. These processes were combined using the operand “OR” in a simple robotic process assessment “BOT”. The presence of any single process marker was termed “BOT positive” or, if none present, “BOT negative”. No other clinical parameters were used. All BOT positive patients who were registered at baseline, were highlighted to their GP, who then validated each case for inclusion or exclusion from the PCR with a binary “yes” or “no” decision. All further clinical considerations arising from this decision, such as contact with the patient or care planning, were a matter for the GP under their routine clinical care. All patients were followed for their mortality status until study end. The study ran for 15 months and a minimum of 6 months after the last recorded GP review.

### Outcomes

Performance metrics, including predictive power for prospective mortality, to assess whether the combined digital and clinical review protocol improved PCR accuracy and uptake.

### Research checklists

Strengthening the Reporting of Observational Studies in Epidemiology for cohort studies was applied to the study protocol (21).

### Data

The data set links demographic, clinical and activity data from primary care, hospital, and community services under General Data Protection Regulation (22,23). The 5 markers were ascertained, frailty was assessed by the Rockwood score, or in its absence the Electronic Frailty Index, and then risk categorised to binary variable of ‘moderate / severe frailty’ versus “non-frail”. Mortality was determined from hospital mortality statistics and rolling NHS Strategic Tracing Service checks.

### Statistical method

All data were analysed on IBM SPPS version 29. Multiple group means were tested by one way analysis of variance, and the Chi-square test was used for the difference between proportions. Performance metrics were determined by receiver operating characterisation and the c-statistic given with its 95% confidence intervals. Binary logistic regression was used to consider the association of independent variables with any categorical dependent variable and Odd’s Ratios (OR) are given with 95% confidence intervals. Survival analysis was by Cox’s regression. Results are presented as the mean ± standard deviation (SD) or as numbers with percentages. Statistical significance of all tests applied was taken at p<0.05.

### Patient consent

Not applicable

### Patient and Public involvement

For this proof-of-concept phase, intended use of integrated clinical data in digital heath care methodologies was discussed with the Trust’s patient liaison group, receiving full support.

## Results

Figure 1. shows the dispersion of the population (n=39,079) by initial PCR category, process marker driven BOT status, and GP assessment. This yielded 4 final groups, **Table 1** gives their demographic, clinical and unscheduled care activity characteristics: 1) not on the palliative care register and BOT negative (n= 38,111, 97.5%); 2), not on the palliative care register, BOT positive, but not added to the PCF by GP review (n=415, 76% of category); 3), not on the PCR, BOT positive, and added to the resister (n=131, 24% of category); 4) already on the PCR (n=422, 1.1%).

**Table 1.** Showing the demographic, clinical and urgent activity characteristics, and prospective mortality outcomes of the 4 groupings derived from the whole population alive at baseline. A/E, Accident and Emergency OR, Odds ratio.

|  | Derived groupings |  |  |  | P value |
| --- | --- | --- | --- | --- | --- |
| Cohort derivation | 1 | 2 | 3 | 4 |  |
| On original palliative care register | No | No | No | Yes |  |
| Any palliative care process marker | None | Present | Present | - |  |
| GP review for addition to palliative care register | Not reviewed | Reviewed “No” | Reviewed “Yes” | - |  |
| Number (total cohort 39,079) | 38,111 (98.9%) | 415 (1.1%) | 131 (0.3%) | 422 (1.1%) |  |
| Age (years) | 47.1 ± 18.0 | 77.0 ± 13.9 | 83.4 ± 12.6 | 80.7 ± 13.2 | P<0.001 |
| Gender (male) | 19,503 (51.2%) | 179 (43.1%) | 53 (40.5%) | 181 (42.9%) | P<0.001 |
| Ethnicity (white) | 21,031 (55.2%) | 354 (85.3%) | 118 (90.1%) | 358 (84.8%) | P<0.001 |
| Index of multiple deprivation | 28.6 ± 16.3 | 23.3 ± 14.0 | 21.0 ± 14.0 | 19.1 ± 11.0 | P<0.001 |
| >=3 A/E attendances in prior 12 months | 437 (1.1%) | 20 (4.8%) | 7 (5.3%) | 30 (7.1%) | P<0.001 |
| >=3 non-elective admissions in 12 prior months | 200 (0.5%) | 35 (8.4%) | 14 (10.7%) | 42 (10%) | P<0.001 |
| >=3 co-morbidities | 3,811 (10.0%) | 297 (71.6%) | 92 (70.2%) | 316 (74.9%) | P<0.001 |
| Frailty (moderate or severe) | 3,863 (10.1%) | 323 (77.8%) | 108 (82.4%) | 349 (82.7%) | P<0.001 |
| Nursing home resident | 104 (0.3%) | 129 (31.1%) | 34 (26.0%) | 231 (54.7%) | P<0.001 |
| Mortality (total deaths 619) | 318 (0.8%) | 81 (19.5%) | 63 (48.1%) | 157 (37.2%) | P<0.001 |
| Mortality OR (95% CI)- unadjusted | 1.0 (comparator) | 28.8 (22.1-37.6) | 110.1 (76.8-88.3) | 70.4 (56.2-88.2) | P<0.001 |
| Mortality OR (95% CI) - adjusted | 1.0 (comparator) | 3.2 (2.3 – 4.3), | 10.1 (6.7 – 15.3) | 6.2 (4.5 – 8.5) | P<0.001 |

**Figure 1.**
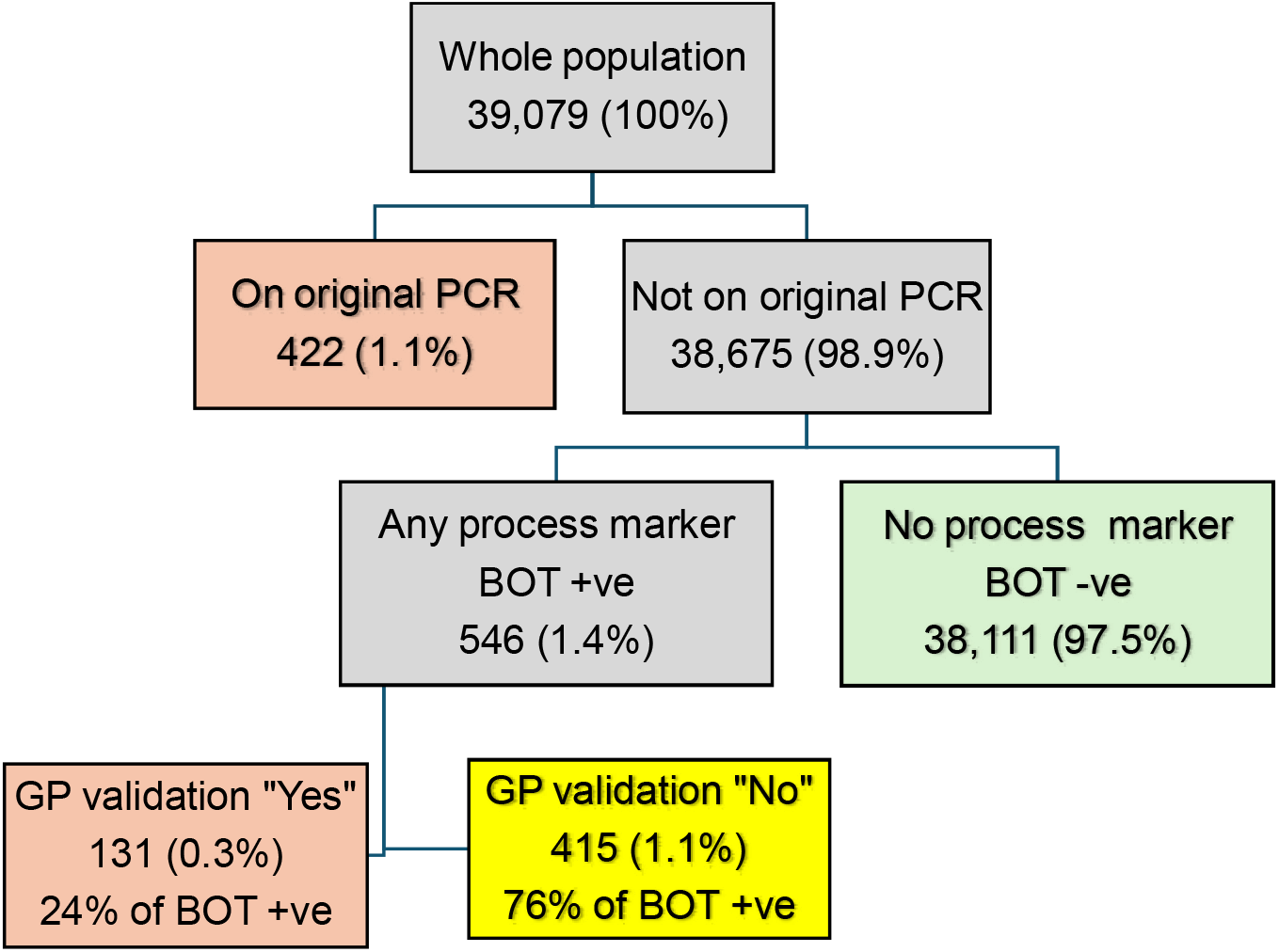
Sequential dispersion of the cohort by initial palliative care registration, the presence of palliative care process markers and clinician review for palliative care registration (PCR).

Figure 2. shows their Cox survival regression. There were 610 deaths over the study duration. Final mortality outcomes differed significantly between the groups. In unadjusted binary logistic regression (*X*^2^ = 1,534.4,p<0.001) of vital status with these groupings, those initially not on the PCR who were “BOT positive” and GP reviewed to join the register had the highest mortality rate and the highest Odds Ratios (OR), compared to those who were not on the register and BOT negative (*X*^2^ = 2,955.8, p<0.001), and also higher than the original PCR (*X*^2^ = 4.9, p<0.05). Those who were not on the register initially and “BOT positive”, but were not added following GP review still had a higher mortality rate than the BOT negative group (*X*^2^ = 1,398.2, p<0.001), but were lower than the original PCR group (*X*^2^ = 32.2, p<0.001). Unadjusted ORs are given in **Table 1**. An adjusted binary logistic regression model of vital status and these groupings, including age (p<0.001), gender (p<0.001), ethnicity (ns), deprivation (p<0.01), unscheduled care (p<0.001), multi-morbidity (p<0.001), frailty (ns) and nursing home residence (p<0.001) remained highly significant (*X*^2^ = 2,458.6, p<0.001) as did the independent 4 group palliative care risk categorisation variable (p<0.001). These adjusted ORs are shown in **Table 1**, showing the differences are independent of demographic and clinical variables.

**Figure 2.**
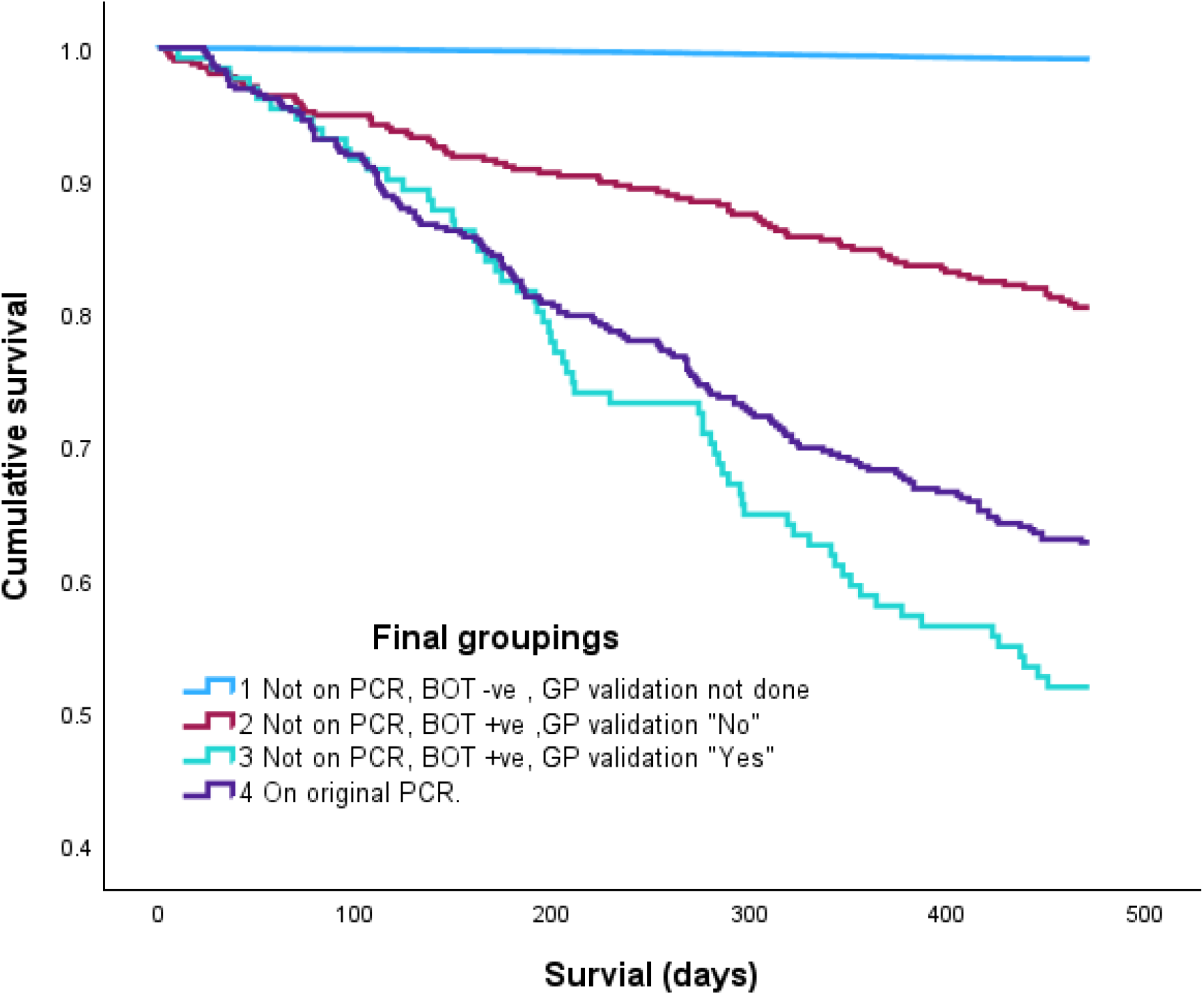
Cox survival regression for the 4 groups as defined in **Table 1**. PCR, palliative care register.

ROC curves (**Figure 3**) show the association of mortality with the original palliative care register group (group 4), the final palliative care register group (combining groups 4 and 3) and a novel digitally derived ‘digital tracker’ grouping which combined all 3 higher risk groups (2, 3, and 4). The original palliative care register captured 157 (32%) of all deaths, the updated revised palliative care register captured 220 (35.5%) whilst the digital tracker identified 301 (48.6%) deaths. The performance metrics of these varying palliative care register constructs are given in **Table 2**. The ROC c-statistics showed sequential improvement which significantly differed from each other (all p<0.001), with particularly marked improvements seen in sensitivity and the positive predictive value.

**Table 2.** Performance metrics of the differing described palliative care register categories for their association with prospective mortality (see Figure 3).

| Palliative care register cohort | Original palliative care register | Final palliative care register | Digital tracker and final palliative care register |
| --- | --- | --- | --- |
| Number (% population) | 422 (1.1%) | 553 (1.4%) | 968 (2.5%) |
| Number of deaths (% total, n = 619) | 157 (25.4%) | 220 (35.5%) | 301 (48.6%) |
| Unadjusted Odds Ratio (95% CI) (all p<0.001) | 49.0 (39.4 - 60.9) | 63.1 (51.9 – 76.9) | 53.6 (45.0 – 63.9) |
| C-statistic (95% CI) | 0.62 (0.61-0.64) | 0.67 (0.66-0.69) | 0.73 (0.72-0.75) |
| Sensitivity - % of those who died identified | 25.4% | 35.5% | 48.6% |
| Specificity | 99.3% | 99.1% | 98.2% |
| Positive predictive value | 29.1% | 36.8% | 41.8% |
| Negative predictive value | 99.2% | 99.1% | 98.6% |
| Accuracy | 98.1% | 98.1% | 97.5% |

**Figure 3.**
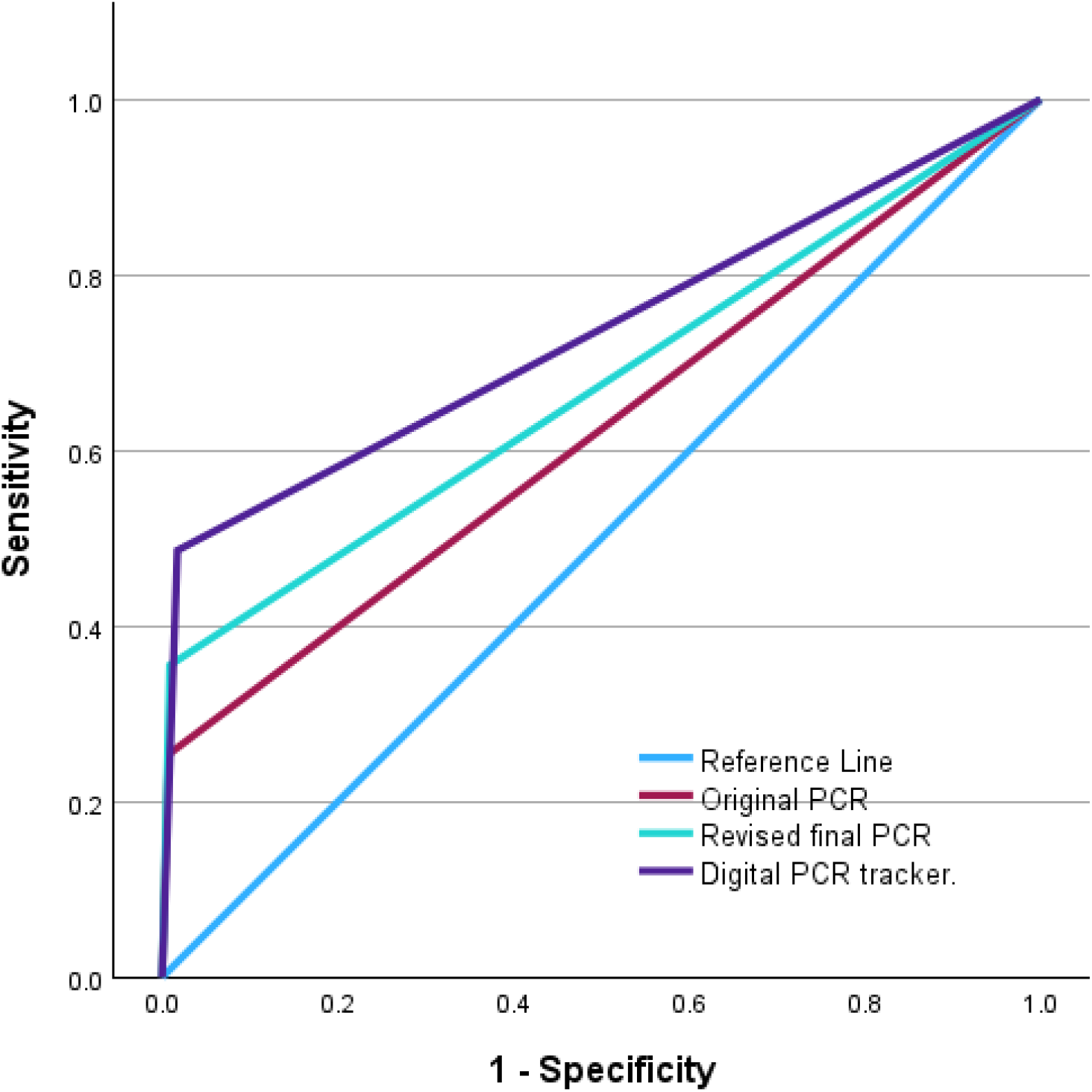
Receiver Operating Curves for the binary association of various palliative care registration (PCR) categories with prospectively ascertained mortality.

## Discussion

### Summary

The robotic process digital tool, combined with GP judgment, escalated 546 patients: 131 (24% with 48.1% 1-year mortality) required PCR; this improved prediction metrics for the enhanced PCR. Those not joined to the PCR (1-year mortality of 19.5%) were retained under surveillance in a “digital tracker” for rolling assessment. This enhanced early identification method is a valid platform for care planning,

### Strengths and limitations

The strengths include a simple methodology, utilising recorded process markers, deployed at large scale, in real-life general practice setting, incorporating clinical judgement. Capturing deaths is important but improving care is the goal, the limitation of our study is that any resulting changes in patient outcomes were not assessed. This is now an important focus of our ongoing research.

### Comparison with existing literature

We are unaware of any prior publications of digital methodologies to specifically improve PCR uptake and accuracy, nor of studies of sequential digital triage followed by clinical validation that improve PCR performance metrics (11). The NHS EARLY Programme(14) adopts the key strategies outlined here: informatics-based risk identification (albeit utilising different variables for the prediction purpose), the incorporation of the Surprise Question to capture clinical judgment, care planning and palliative care registration. However, NHS EARLY has not published its metrics or outcomes. Other mortality prediction engines for earlier identification have been developed in small, often disease specific cohorts, their performance metrics are uncertain, they are generally not automated, they have not been deployed or studied at a whole population scale (17,24) and none incorporate the outcome of actual PCR, or track prospective outcomes, or have built in rolling review.

### General discussion

Care at the end of life affords limited time to get care planning and delivery right. Too many patients whose deaths are deemed expected remain missing from PCR (7,8) There is a dearth of evidence regarding failed ascertainment (25). Identifying patients for PCR must have accuracy in predicting mortality, but such evidence for currently recommended strategies for identifying patients for PCR is scant (26).

Our straightforward digital methodology demonstrates a replicable, whole population approach which improves PCR identification, leading to significantly improved performance metrics of the final PCR for deaths at 1 year, a strong affirmation of the selection’s validity. We emphasise that identification of patients at risk of dying in the next 12 months who could benefit from end-of-life care should be seen as a distinct task. PCR inclusion is not solely about mortality but a shifting approach towards holistic needs assessment with a focus on quality of life, meaning that patients may be on a PCR for several years.

Our method is distinct in 3 key aspects. Firstly, we have extensively evaluated the variables we use (19,20). Many single variables have standalone statistical association with mortality, but in multiple variable use, they become non-significant due to collinearity, for example, counterintuitively, the electronic frailty index does not independently predict mortality in our systems, and thus the possible need for end-of-life registration. Crucially, compared to other methodologies, our system is disease agnostic, which is important when considering multimorbidity. We have published effective mortality prediction models (20,27), however they predict a future which may or may not happen. Here, we use robotic process not prediction, describing the “here and now”, counting what has already been done, highlighting what has probably been missed, thus creating a live clinical governance and quality improvement tool for clinician use in direct care.

Secondly, we reaffirm(28), that GP clinical judgment improves uptake and accuracy beyond data mechanisms. Clinical judgment within the system becomes a process measure of its own right (29), clearly benefiting cohort definition, now validated by prospective mortality prediction and allaying reservations about clinician identification of those nearing end-of-life (30). Notably clinical judgement also led to exclusion from PCR of BOT-positive patients (415/546 (76%, mortality 19.5%). Register inclusion requires greater consideration than mortality expectation (4). The reasons for this were not recorded but will be multiple and complex: “grey case” uncertainty, clinical complexity, the need for difficult conversations, assessment of palliative care needs, patient choice and consent, engagement and misjudgement.

Thirdly, we address the above in two ways. The PCR excluded BOT +ve group were retained in a digital safety net, the ‘digital tracker’, which captured 48% of deaths at 1 year. With the use of the digital tracker, this system improves PCR quality but may also push the ambition of early detection and anticipatory care planning into this “grey zone”. Our original PCR was already 1.1% (422, capturing n=157 (25.5%) deaths), whereas the tracker encompassed 2.5% (n=968, 48% of deaths), well beyond the targeted 1% registration concept. Furthermore, PRADA is used continuously, not as a one off or at set intervals. If a patient’s circumstances change, clinicians are prompted to reassess. This iterative approach also captures newly identified patients. We emphasize that under no circumstances do we propose this as a digital replacement for the palliative care register, and that no automated interventions must arise out of this clinical decision support aid and surveillance tool. It must be highlighted that even with the ability to improve early identification that outcomes will not improve unless there are effective mechanisms of needs analysis and care planning in place.

### Implications for research and practice

Our future research will address this gap, exploring whether this method leads to more timely needs analysis, care planning, reduced urgent care and deaths in hospital. Critically, we will also explore the views of patients and their families including perspectives on the quality of care they receive.

Policy makers, commissioners and care providers must be challenged to adopt innovation early into practice. Our described digitised palliative care system is easily replicable, uses rudimentary digital techniques, is applicable at scale, within primary care electronic systems, in a manner that is amenable to quality measurement and clinical governance, including rolling assessment of outcomes and learned system improvements. This must be actioned now, there is an NHS opportunity with NHS EARLY (also a digital robotic process system), to deploy these emergent methods rapidly, without the inertia of waiting for machine learning approaches, already 20 years in gestation.

Research funding must enable the research community to verify, refute or enhance this first ever demonstration of a digital health function to transform the quality and uptake of palliative care registers and explore the intended benefits beyond increased PCR accuracy and mortality prediction.

## Conclusion

This novel digital system gives oversight of the PCR to services and clinicians in a live front-line care overview, identifying those potentially requiring PCR and those in whom registration is not timely but still of higher mortality risk. This facilitates a shift away from opportunistic interventions towards proactive identification and anticipatory care planning, and we propose it as a step change in care at the end-of-life.

## Supporting information

STROBE Checklist

## Data Availability

All data produced in the present study are available upon reasonable request to the authors

## Abbreviations Used

BOT: automated robotic process
GP: General Practitioner
OR: Odds Ratios
PCR: Palliative care register
PRADA: Proactive Risk-Based and Data-Driven Assessment of Patients at the End of Life
SD: Standard deviation

## Additional Information

### Ethical Approval

The study did not involve selection criteria, randomisation, nor any intervention that ought not to have otherwise happened. Our methodology was discussed with our Institutional Review Board (Royal Wolverhampton NHS Trust Research and Development Department) who deemed that ethical approval and informed consent was not necessary for this study. The study was considered a clinical governance driven quality improvement project using innovative digital healthcare methodologies.

## Acknowledgements

None.

## Data Sharing

The corresponding author will share anonymised study data upon reasonable request, subject to approval of any proposal by the authors and our research governance body.

## Copyright

The corresponding author has the right to grant on behalf of all authors, the license to permit this article to be published.

## ±Supplementary Data

